# Comparison of LA and PVC mapping using OCTARAY and OPTRELL catheters

**DOI:** 10.1101/2025.03.03.25323279

**Authors:** Jumpei Saito, Kato Daiki, Sato Hirotoshi, Toshihiko Matsuda, Yui Koyanagi, Katsuya Yoshihiro, Yuma Gibo, Soichiro Usumoto, Taro Kimura, Suguru Shimazu, Wataru Igawa, Seitaro Ebara, Toshitaka Okabe, Naoei Isomura, Masahiko Ochiai

## Abstract

**Background:** Multielectrode mapping catheters, such as the OCTARAY and OPTRELL, are essential in creating myocardial electroanatomical mapping in arrhythmias. The OCTARAY is a multi-spline mapping catheter with 48 closely spaced multielectrodes that enables high-resolution electroanatomical mapping, while the OPTRELL is a multi-electrode catheter with 36 electrodes arranged on eight radiating splines. However, only a few studies have compared their performance. In this study, we aimed to compare the OCTARAY and OPTRELL catheters in two areas: left atrial (LA) mapping during atrial fibrillation (AF) ablation; and premature ventricular contraction (PVC) mapping.

**Methods:** Twenty patients (Ten patients for LA mapping and ten for PVC mapping) were enrolled. LA voltage mapping was performed twice, alternating between catheters post-AF ablation. Parameters compared included mapping time, mapping points, catheter-induced premature atrial contraction (PACs), tissue proximity indication, low voltage area, and fluoroscopy time. For PVC mapping, comparisons included mapping time, catheter-induced PVCs, earliest activation time measured from the onset of PVC QRS, earliest activation point, and fluoroscopy time.

**Results:** Compared with THE OCTARAY, mean voltage using the OPTRELL was higher (0.192 mV[0.072, 0.48] vs. 0.126 mV[0.042, 0.378]; *P* = .001) and the percentage of tissue proximity indication positive was also higher (14.97% vs. 11.45%; *P* < .001). However, there were no significant differences in low voltage area between the two groups (39.5 m^2^[16.5, 66.8] vs. 40[23.6, 61]; *P* = .861), and in other LA parameters. In PVC mapping, catheter-induced PVCs using OPTRELL were significantly fewer than the OCTARAY (100 [32, 337] vs. 247 [110, 745], *P* = .039), with fewer catheter induced PVCs per minute (15 [6, 23] vs. 35 [20, 71], *P* = .039). However, no significant differences were observed in other PVCs mapping parameters.

**Conclusion:** The OPTRELL catheter demonstrated higher voltage recordings in LA mapping and fewer catheter-induced PVCs compared with the OCTARAY catheter. However, no significant difference was observed in other mapping parameters.

## INTRODUCTION

Multielectrode mapping catheters play a crucial role in creating myocardial electroanatomical mapping for all types of arrhythmias. However, the accuracy of intracardiac mapping is significantly influenced by several factors, including tissue contact, mapping catheter characteristics (such as electrode size, spacing between electrodes, and orientation relative to the tissue). In addition, while there are several multielectrode mapping catheters available, only a few studies compare performance of different multi-electrode mapping catheters. The OCTARAY multielectrode mapping catheter (Biosense Webster, Diamond Bar, CA, USA) is an advanced iteration of the PENTARAY catheter (Biosense Webster, Diamond Bar, CA, USA). THE OCTARAY consists of eight splines, each with six electrodes measuring 0.5 mm width (0.9 mm^2^ surface area) and spaced 2 mm apart center-to-center, as shown in Figure 1A. Previous studies have demonstrated that the OCTARAY records more electrograms (EGMs) per map in a shorter mapping time, resulting in higher point density and a faster point acquisition rate compared with the PENTARAY (1, 2). The OPTRELL multielectrode mapping catheter (Biosense Webster, Diamond Bar, CA, USA) features 36 electrodes (each with a surface area of 0.9 mm^2^) distributed across six splines (six electrodes per spline), with a symmetric center-to-center interelectrode distance of 2.4 mm along and across the splines, as shown in Figure 1B.

**Figure 1.**
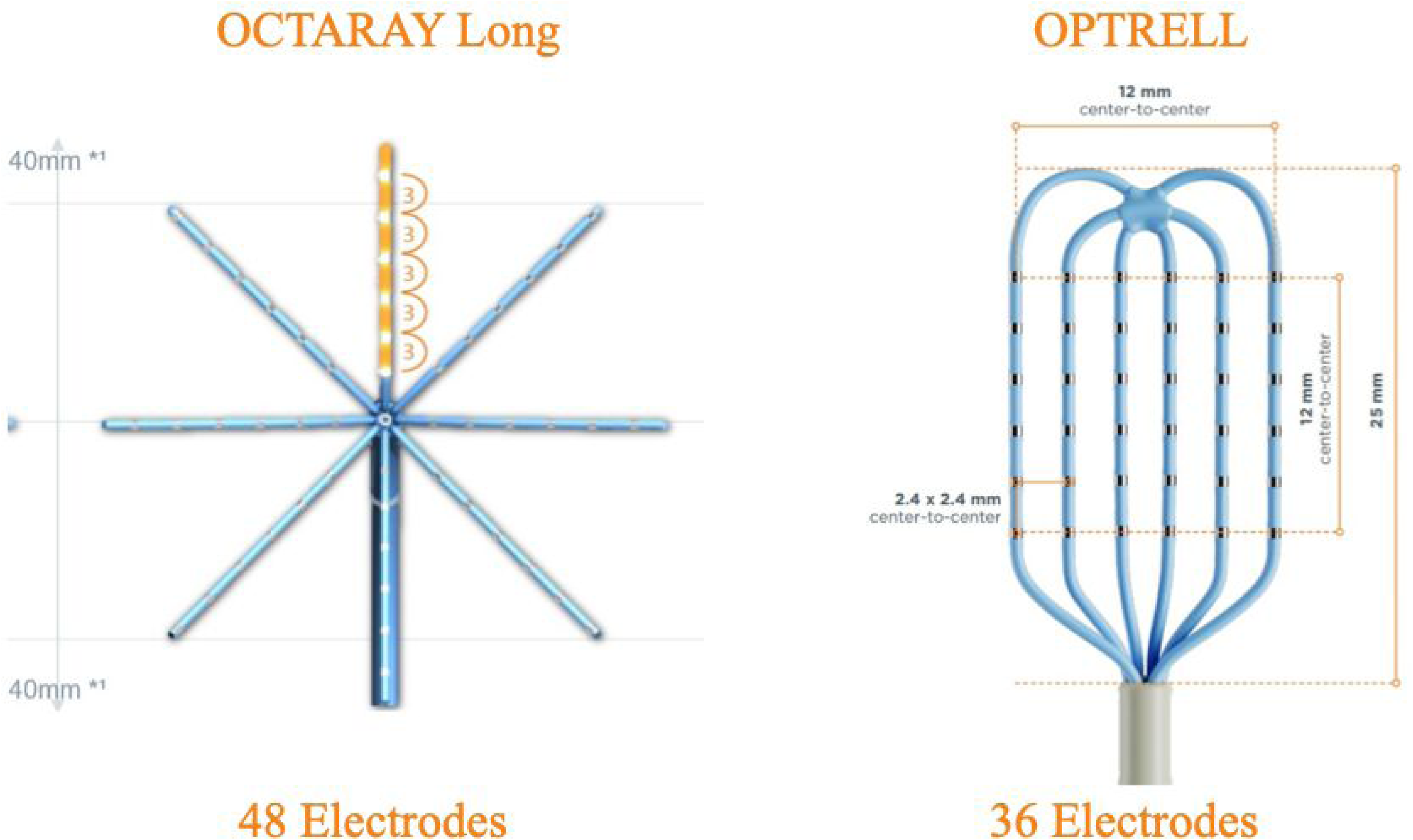
The OCTARAY Long 3-3-3-3-3 catheter and the OPTRELL catheter. Catheter shape, distribution of electrodes, and measurement of each electrode.

Left atrial (LA) voltage mapping is performed to identify the arrhythmogenic substrate of the atria. Areas of reduced bipolar voltage are defined as low-voltage areas (LVAs) and correspond to areas of fibrosis (3–5). After pulmonary vein isolation (PVI), LA LVAs have been identified as predictors of atrial fibrillation (AF) recurrence and long-term incidences of heart failure, stroke, and death (6). Previous studies have shown that LVAs ablation can reduce the risk of AF (7–10). Therefore, it is important to assess the extent of LVAs.

Frequent premature ventricular contractions (PVCs) can cause significant symptoms and are associated with an increased risk of congestive heart failure and mortality in the general population (11, 12). Catheter ablation for PVCs has been shown to restore left ventricular function in patients with PVC-induced cardiomyopathy (13–16), and to decrease B-type natriuretic peptide (BNP) levels while increasing estimated glomerular filtration rate even in patients with preserved left ventricular ejection fraction (LVEF) (17). Intracardiac mapping plays an important role in PVC ablation by accurately identifying the site of PVCs, thereby increasing the success rate of ablation. There were two purposes of this study. First, we aimed to compare electroanatomical LA mapping between the OCTARAY and the OPTRELL catheter in patients who underwent AF ablation and assess LA mapping parameters. Second, we compared PVC mapping parameters with the OCTARAY and OPTRELL catheters in patients who underwent PVC ablation (Figure 2).

**Figure 2.**
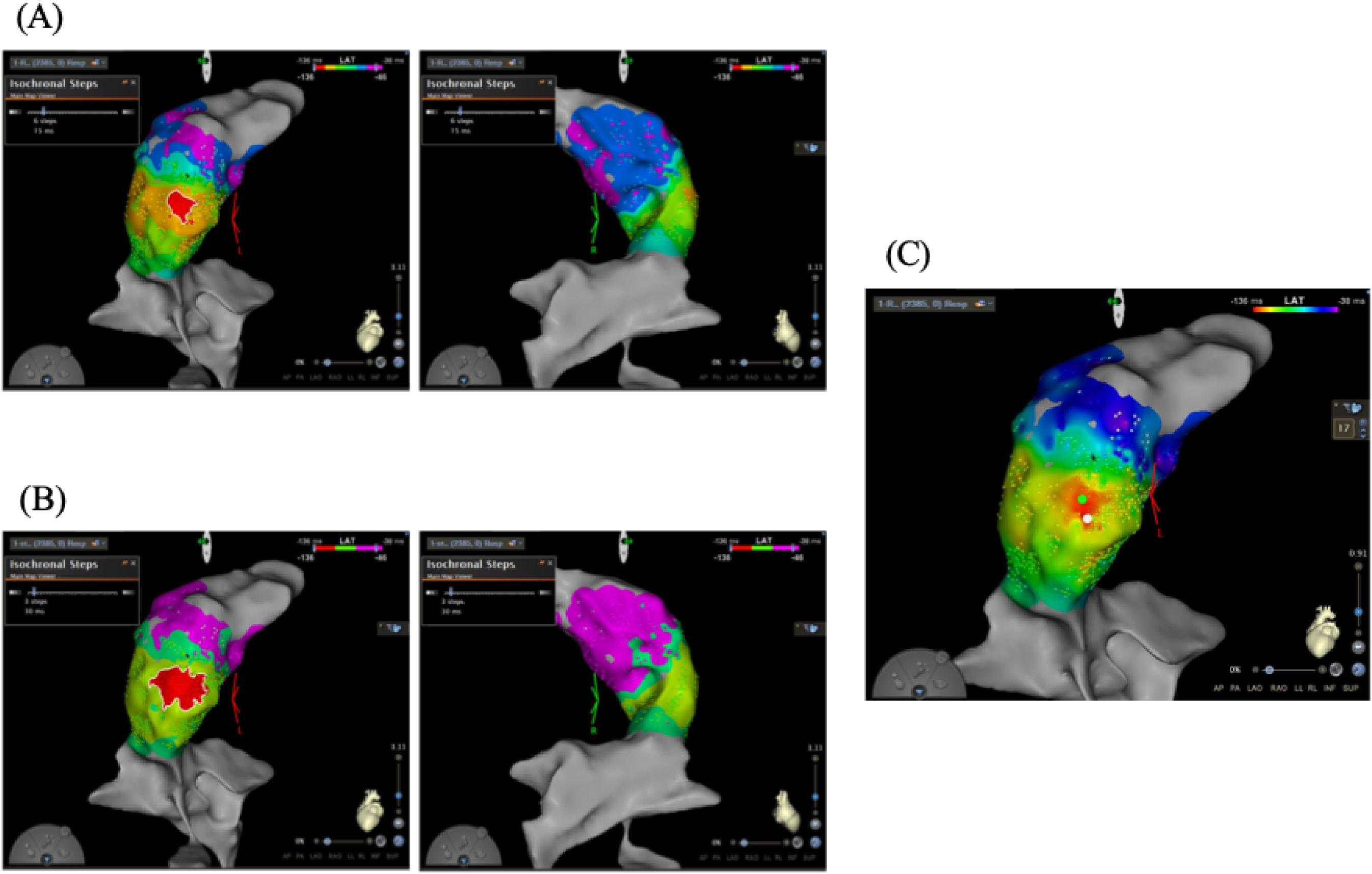
Representative cases of premature ventricular contraction (PVC) originating at the right ventricular outflow tract (RVOT). (A) The red area shows 15 msec isochronal area, measuring 0.8 cm^2^. (B) The red area shows 30-msec isochronal area, measuring 1.9 cm^2^, which is larger than the 15-msec isochronal area. (C) This figure shows the white tag as the success point and the blue tag as the earliest activation site (EAS). The distance from the white tag to the blue tag is 3.4 mm.

## METHODS

### Patients

A total of twenty consecutive patients (ten for left atrial mapping and another ten for PVC mapping) were enrolled between May and December 2024. Exclusion criteria included: age <20 years, prior cardiac surgery, pacemaker implantation, patients in whom sinus rhythm was unsustainable during voltage mapping in AF ablation, and patients in whom PVCs did not occur frequently enough to allow for effective mapping during PVC ablation. This study complied with the principles of the Declaration of Helsinki. Written informed consent for the ablation procedure and participation in the study was obtained from all patients, and the protocol was approved by our Institutional Review Board.

### Procedure of AF ablation

Electrophysiological studies and catheter ablation were performed under deep sedation. A 6-French (Fr) decapolar electrode was placed in the coronary sinus. Following a transseptal puncture in the fossa ovalis, a VIZIGO (Biosense Webster) and SL0 (Abbott) sheath were introduced into the LA using the single transseptal puncture technique. An ablation catheter (SMART TOUCH or Q-DOT; Biosense Webster) and a 20-pole circular mapping catheter (Lasso catheter; Biosense Webster) were inserted into the LA using the SL0 sheath. Thereafter, voltage mapping using either OCTARAY or OPTRELL catheter and ablation were performed under the guidance of the CARTO three-dimensional mapping system (Biosense Webster). Intravenous heparin was administered to maintain an activated clotting time (ACT) of 300 s throughout the procedure. First-pass encirclement was performed using a point-by-point ablation approach with an ablation index (AI)-guided 40–50 W power ablation protocol (AI anterior: 450; AI posterior: 400). Catheter operators aimed to space lesions less than 5 mm apart. Additional ablation techniques, including LVA homogenization, linear ablation, and ablation for non-pulmonary vein AF triggers, were incorporated at the discretion of the attending physicians following LA mapping.

### LA voltage mapping

After achieving PVI, voltage mapping under high right atrial pacing at 600 msec was performed twice using each catheter (half of the cases used the OCTARAY first, and the remaining cases used the OPTRELL first). The mapping catheters were inserted into the LA through the VIZIGO sheath to facilitate better catheter-tissue contact and stability. Adequate catheter contact with the atrial wall was confirmed by fluoroscopic guidance when necessary. The CARTO system’s default filter settings were used for recording bipolar signals, including a 16-Hz high-pass filter and a 500-Hz low-pass filter. Similarly, unipolar signals were recorded between each electrode of the mapping catheter or the distal tip of the ablation catheter and the Wilson central terminal, with a filter setting of 2–240 Hz. Furthermore, a notch filter was applied to eliminate noise from the environment power lines. Mapping points were automatically acquired using the following criteria: cycle length stability within ±20 msec, local activation time stability within 4 ms, catheter position stability of 4 mm/sec, and point density of 1mm. During voltage mapping, the following parameters were recorded: (1) mapping time, (2) number of mapping points, (3) LA surface area, (4) LA volume, (5) tissue proximity indication (TPI), (6) number of catheter-induced premature atrial contractions (PACs), and (7) fluoroscopy time. After mapping, all electrograms were manually reviewed to exclude noise or artifacts.

The LVA size was manually measured on each voltage map. LVA was defined as a region with a bipolar voltage of <0.50, the border zone area was defined as a region with a bipolar voltage of <1.0, or <1.5 mV.

### Procedure of PVC Ablation

Electrophysiological studies and catheter ablation were performed under deep sedation with intravenous propofol. A 6-Fr linear electrode catheter was placed in the coronary sinus as far as possible. The initial preferred vascular access was chosen primarily on the morphology of the PVC as observed on the 12-lead electrocardiogram and at the discretion of the investigator. For PVCs of left ventricular origin that could not be evaluated by retrograde access via the aorta, a single transseptal puncture was performed under fluoroscopy in conjunction with intracardiac ultrasound. Intravenous heparin was administered prior to transseptal puncture or retrograde access to maintain an ACT of 300 to 400 seconds. PVC mapping was performed twice using each catheter (half of the cases used the OCTARAY first, and the remaining cases used the OPTRELL first). The mapping catheters were inserted into the ventricle through the VIZIGO sheath to facilitate better catheter-tissue contact and stability.

### PVC mapping details

During voltage mapping, the following mapping efficacy parameters were recorded: (1) mapping time, (2) number of mapping points, (3) number of accepted clinical PVCs, (4) number of mapping catheter-induced PVCs, (5) distance from the earliest activation site (EAS) to the successful PVC ablation point, (6) earliest local ventricular activation timing relative to QRS onset, (7) fluoroscopy time and dose, and (8) 15- and 30-msec isochronal areas.

Mapping time was defined as the time from positioning the mapping catheter near the site of origin of PVC to the point where the operator could determine that the EAS had been identified. The number of accepted PVCs and mapping catheter-induced PVCs were counted by the operator after the procedure. Local activation time (LAT) annotation was performed automatically by the CARTO three-dimensional mapping system, which uses the maximum negative slope of the unipolar distal signal to set the timing of the mapping annotation. After mapping, all electrograms were manually reviewed to exclude noise or artifacts. The end of mapping was determined by the surgeon when the EAS was identified. Isochronal areas defined as the regions surrounded by the propagated wavefront on the PVC maps at 15 and 30 msec after the earliest right ventricular activation. Representative cases demonstrating these mapping parameters are shown in Figure 2. PVC ablation was performed using Q-DOT catheter (Biosense Webster, Diamond Bar, CA, USA) with a power setting of 30–50 W, an irrigation flow rate of 4–15 mL/min, a temperature limit of 45 °C, and a contact force >5g. With reduced (<1 PVC/min) or absent PVC, intravenous administration of isoproterenol and programmed ventricular stimulation were performed. If the number of PVCs remained too low to allow for effective PVC mapping, the patient was excluded from the study.

### Statistical analysis

Continuous data are expressed as median (interquartile range). Categorical data are presented as absolute values and percentages. Differences in variables between the OCTARAY and the OPTRELL were analyzed using the Wilcoxon signed-rank test for continuous variables, and the chi-square test or Fisher’s exact test for categorical variables. Pearson’s correlation coefficient analysis was performed to assess correlations between continuous variables. All analyses were performed using the commercial software (JMP).

## RESULTS

### Patient characteristics

Patient characteristics and clinical outcomes are shown in Table 1 (% male, age ± years, mean LVEF ± %). LA voltage mappings using the OCTARAY and the OPTRELL were completed in all patients with AF without any complications. All patients with PVC underwent successful PVC ablation, and there were no serious complications.

**Table 1.** Baseline patient characteristics.

| characteristics | LA mapping group (N=10) | PVC mapping group (N=10) |
| --- | --- | --- |
| Age, y | 79 (75.5,81.5) | (51,80) |
| Male sex, n (%) | 5 (50) | (75) |
| Body mass index, kg/m<sup>2</sup> | 23.2 (20.0, 27.8) | (20.1, 32.2) |
| CHAD<sup>2</sup>DS<sup>2</sup>-VASc score | 4 (4, 5.3) | - |
| Hypertention, n (%) | 9 (90) | (23.5) |
| Heart failure, n (%) | 4 (40) | (23.5) |
| RV PVC, n (%) | - | 6 (60) |
| LVEF, % | 59 (50, 65.5) | (41.2, 68.4) |
| LAVI, ml/m<sup>3</sup> | 46 (35.3, 56) | (20.5, 64) |

Data are presented as n (%) or median (interquartile range).
Abbreviations: CHA<sup>2</sup>DS<sup>2</sup>-VASc, congestive heart failure, hypertension, age $\geq 75$ years (doubled), diabetes mellitus, prior stroke or transient ischemic attack or thromboembolism (doubled), vascular disease, age 65–74 years; RV, right ventricle; PVC, premature ventricular complex; LVEF, left ventricular ejection fraction; LAVI, left atrial volume index

### LA voltage mapping

The mean voltage by the OPTRELL (0.192 mV [0.072, 0.48]) was higher than that recorded by the OCTARAY (0.126 mV [0.042, 0.378], p = .001) across the entire LA. Specifically, the mean voltage recorded by the OPTRELL was higher in the septal, posterior, roof, and bottom regions compared to that recorded by the OCTARAY (Figure 3). The sizes of LVAs and border zone areas are compared in Figure 4. No significant differences were observed in any region of the LA. Mapping points using the OPTRELL were significantly greater than those using the OCTARAY; however, there was no significant difference in mapping points per minute between the two mapping catheters (Table 2). The percentage of TPI positivity was significantly higher with the OPTRELL. There were no significant differences in other mapping parameters.

**Figure 3.**
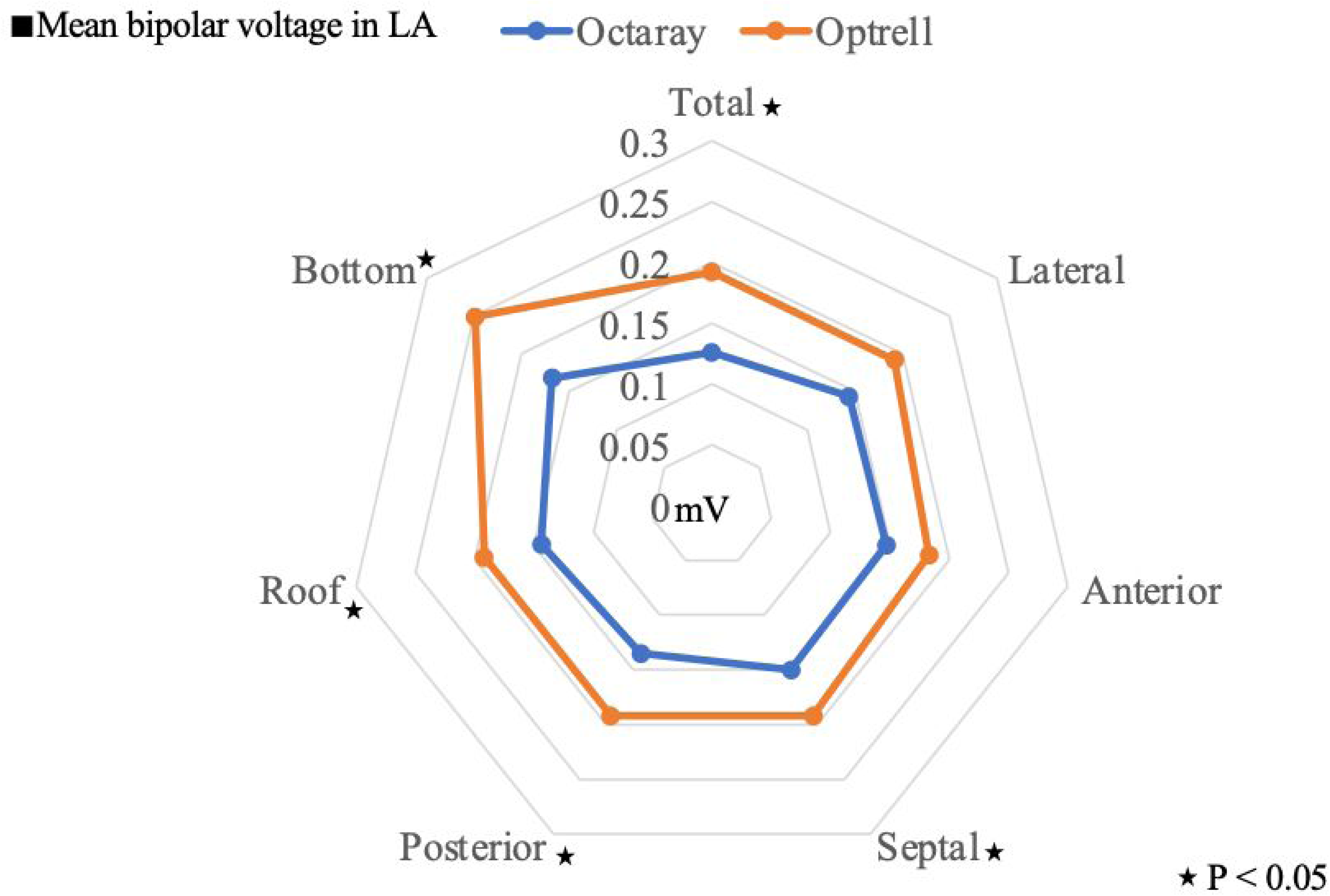
The radar chart indicates mean regional voltages. The OPTRELL demonstrates higher mean voltage.

**Figure 4.**
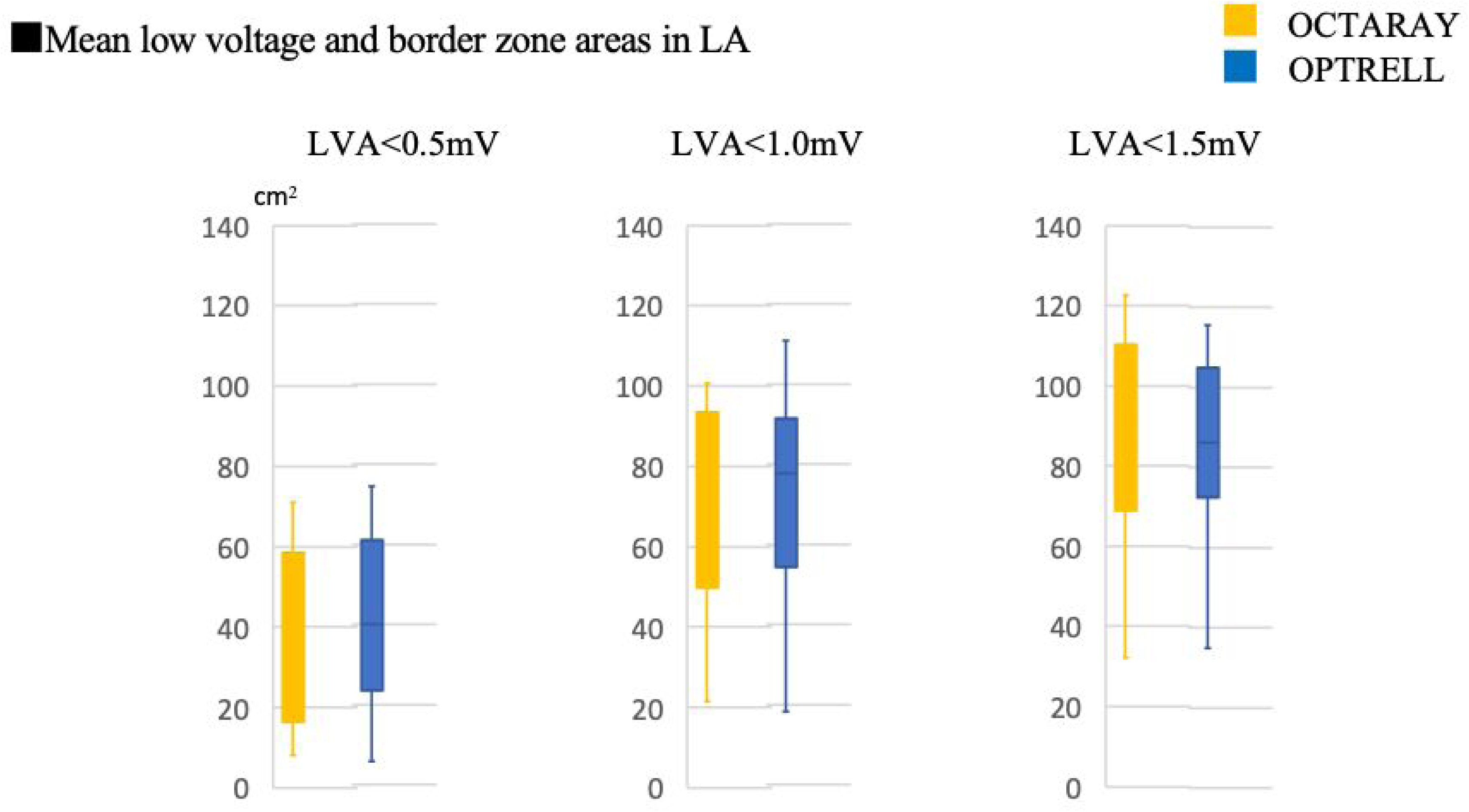
Comparisons of low-voltage and border zone areas. There were no significant differences in low-voltage and border zone areas.

**Table 2.**
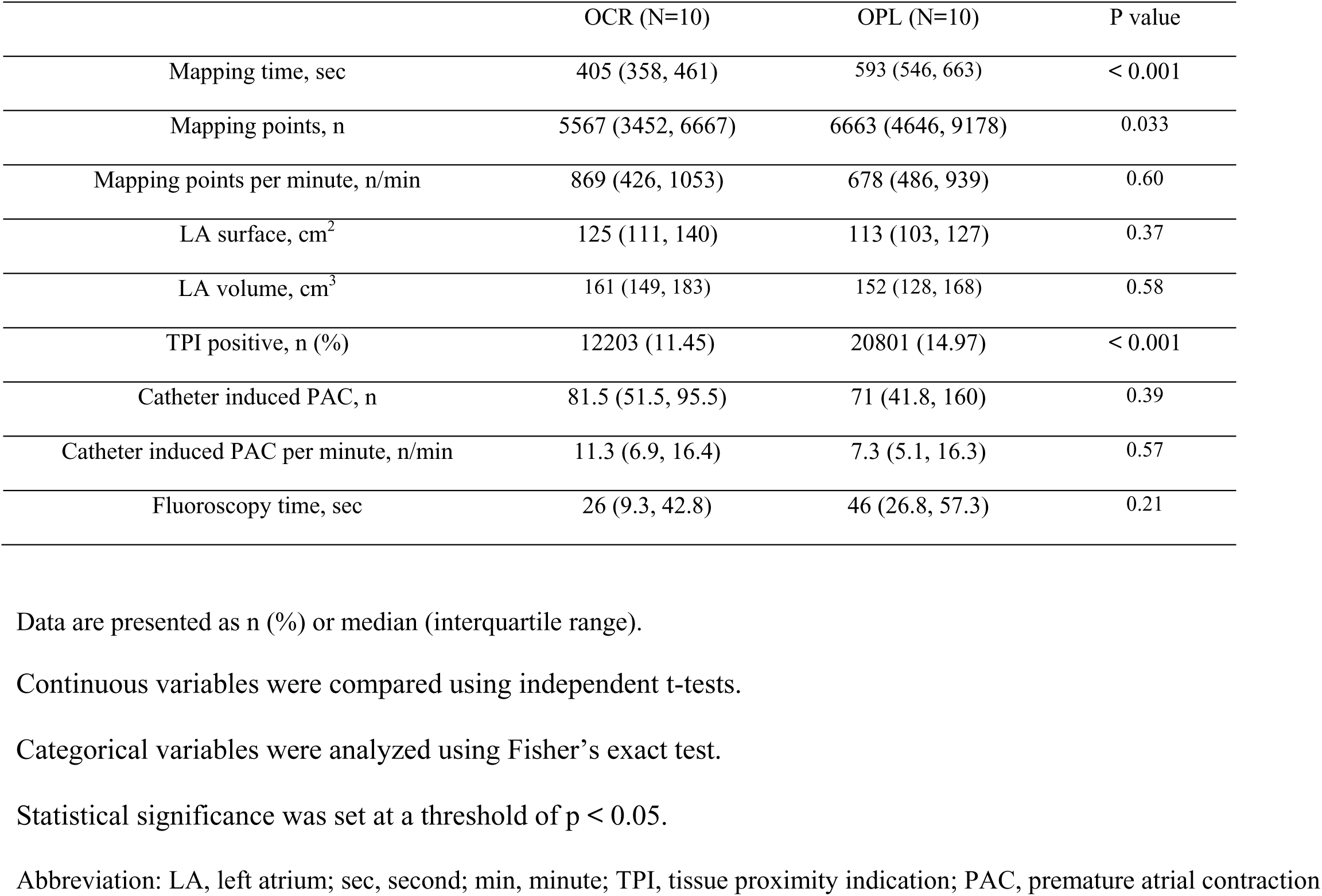
LA mapping parameter.

### PVC mapping

Table 3 shows the PVC mapping parameters. The number of mapping catheter-induced PVCs with the OPTRELL was significantly less than that with the OCTARAY (100 [32, 337] vs. 247 [110, 745], *P* = .039). The number of mapping catheter induced PVCs per minute with the OPTRELL was also lower than that with the OCTARAY (15 [6, 23] vs. 35 [20, 71], *P* = .039). There were no significant differences in other PVC mapping parameters.

**Table 3.**
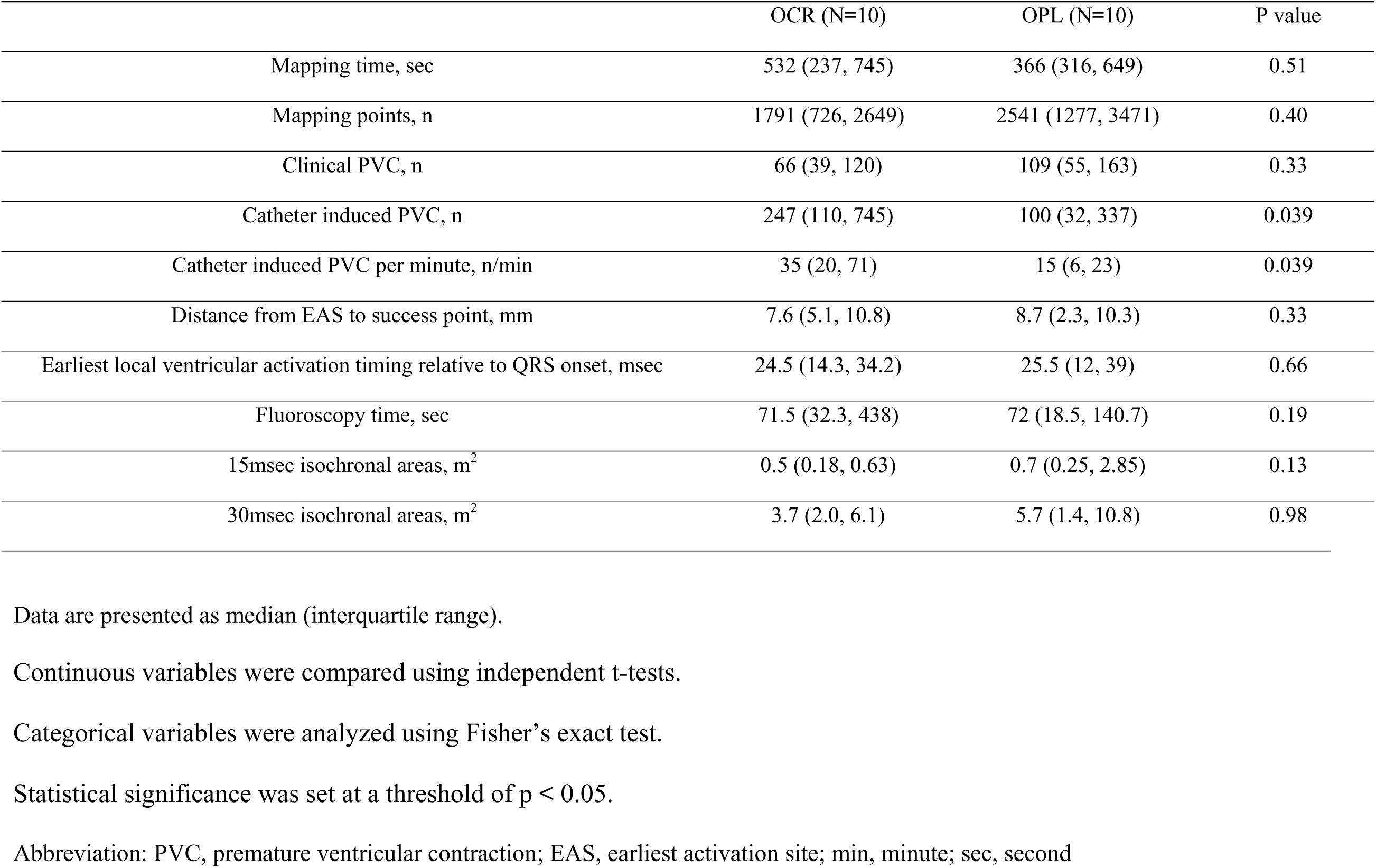
PVC mapping parameter.

## DISCUSSION

This observational study investigated LA and PVC mapping in 10 patients each, comparing the OCTARAY catheter and the OPTRELL catheter. The major findings were as follows: (1) The mean bipolar voltage in LA mapping was higher for the OPTRELL than for the OCTARAY, (2) The number of mapping catheter-induced PVCs and the number per minute with the OPTRELL were significantly lower than with the OCTARAY in PVC mapping.

### LA voltage mapping

The LA voltage map obtained with the OPTRELL catheter demonstrated a 52.4% greater mean bipolar voltage than the OCTARAY. Several studies have compared multielectrode mapping catheters in LA voltage mapping. A previous study showed that the OCTARAY recorded more EGMs per map in a shorter mapping time, resulting in higher point density and a faster point acquisition rate compared with the PENTARAY (1, 2). The HD GRID (Abbott, St. Paul, MN, USA), which has a design similar to the OPTRELL, recorded more mapping points and identified a smaller low-voltage area than the PENTARAY (18). However, no study has compared the OCTARAY and the OPTRELL; thus, our study is the first to do so. In addition, the percentage of TPI positivity was significantly higher with the OPTRELL. TPI uses impedance changes to indicate tissue contact. The impedance of an electrode in blood varies on its location in the heart, so a base impedance map is created to normalize these values. The base impedance map is a three-dimensional function that estimates electrode impedance based on its surface geometry and is generated through a process known as base impedance map training. When an electrode makes contact with tissue, the impedance ratio compared with the base impedance map is calculated and compared with a 7% threshold. A rapid change in impedance of at least 7% from the baseline (blood pool impedance) defines the electrode as TPI positive. Okumura et al. highlighted the importance of assessing catheter-tissue contact using TPI (19, 20). The higher mean LA voltage measured with the OPTRELL compared to the OCTARAY may be due to improved catheter-tissue contact. The OPTRELL’s rectangular shape allows its electrodes to make extensive and consistent contact with the atrial endocardium. In contrast, some of the OCTARAY’s electrodes may not maintain consistent contact with the endocardium, potentially resulting in poorer signal recordings.

### PVC mapping

Intracardiac mapping is important in the ablation of PVCs to accurately identify the site of origin and increase the success rate of ablation (21–24). However, few studies have compared multielectrode mapping catheters in PVC mapping. Our study is the first to compare PVC mapping using multielectrode mapping catheters. The number of mapping catheter-induced PVCs and the number per minute were significantly lower with the OPTRELL than with the OCTARAY in PVC mapping. We hypothesize that the OPTRELL induced fewer PVCs than the OCTARAY due to its ability to engage with the tissue in a more planar manner, thereby distributing contact pressure more evenly. The OCTARAY tended to require longer mapping times and resulted in fewer mapping points than the OPTRELL in PVC mapping. PVC mapping with the OCTARAY may be more challenging due to the higher incidence of catheter induced PVCs, which could lead to longer mapping times and fewer mapping points. However, there was no significant differences in other PVC mapping parameters, indicating that PVC mapping could be performed effectively with the OCTARAY compared to the OPTRELL.

### Limitation

There were some limitations to this study. First, the results represent a single-center experience and may be dependent on specific operators’ skills. Second, the statistical analyses were potentially influenced by the relatively small size of the study population. Consequently, multicenter studies involving larger populations are imperative to establish reliable conclusions regarding the general applicability of this method. Third, the selected mapping points may have been insufficient. Some areas might have been missed despite efforts to achieve even contact with the endocardium. Furthermore, Sekihara et al. demonstrated that the substrates of AF can vary with pacing cycle length (25). Therefore, electroanatomical mapping could be affected by many factors, and it remains unclear whether the original potentials are reliably recorded. Finally, the origin of PVCs estimated by the response to the radiofrequency application might be subject to variability. This is because conductive heating from radiofrequency application can injure adjunctive tissue near the ablation site.

## CONCLUSIONS

The OPTRELL catheter demonstrated higher voltage recordings in LA mapping and fewer catheter-induced PVCs than the OCTARAY catheter. There were no significant differences in other mapping parameters.

## Data availability statement

The original contributions presented in the study are included in the article, and further inquiries can be directed to the corresponding author.

## Ethics approval statement

This study involved human participants and was approved by the Showa University Ethics Committee (ID: 2024-288-B).

## Patient consent statement

The patients provided written informed consent to participate in this study.

## Abbreviation

LA: Left Atrium
PVC: Premature Ventricular Contraction
AF: Atrial Fibrillation
EGM: Electrogram
TPI: Tissue Proximity Indication
LVA: Low-Voltage Area
PVI: Pulmonary Vein Isolation
ACT: Activated Clotting Time
AI: Ablation Index
CF: Contact Force

## PERSPECTIVES

## COMPETENCY IN MEDICAL KNOWLEDGE

The OPTRELL catheter demonstrates superior performance in left atrial (LA) voltage mapping and PVC mapping, with higher voltage recordings, fewer catheter-induced PVCs, and improved tissue contact compared to the OCTARAY. These findings suggest that the OPTRELL may enhance the accuracy and efficiency of electroanatomical mapping, leading to better identification of arrhythmogenic substrates and improved ablation outcomes.

## TRANSLATIONAL OUTLOOK

Barriers to clinical translation include operator dependency, small study populations, and potential variability in arrhythmia substrates. Future research should focus on multicenter studies, advanced mapping technologies, and cost-effectiveness analyses to validate the OPTRELL’s benefits and facilitate its broader adoption in clinical practice.

